# Identification of consensus head and neck cancer-associated microbiota signatures: a meta-analysis of 16S rRNA and The Cancer Microbiome Atlas datasets

**DOI:** 10.1101/2023.07.25.23293137

**Authors:** Kenny Yeo, Runhao Li, Fangmeinuo Wu, George Bouras, Linh T.H. Mai, Eric Smith, Peter-John Wormald, Rowan Valentine, Alkis James Psaltis, Sarah Vreugde, Kevin Fenix

## Abstract

**Objective:** Multiple reports have attempted to describe the tumour microbiota in head and neck cancer. However, these have failed to produce a consistent microbiota signature which may undermine understanding the importance of bacterial-mediated effects in head and neck cancer. The aim of this study is to consolidate these datasets and identify a consensus microbiota signature in head and neck cancer.

**Methods:** We analysed 11 published head and neck cancer 16S ribosomal RNA microbial datasets collected from cancer, cancer-adjacent and non-cancer tissue to generate a consensus microbiota signature. These signatures were then validated using The Cancer Microbiome Atlas database.

**Results:** We identified unique bacteria enrichment within tissue types and correlated it with possible functional and clinical outcomes.

**Conclusions:** Our meta-analysis demonstrates a consensus microbiota signature for head and neck cancer, highlighting its potential importance in this disease.

**Highlights:**

- The first meta-analysis of tissue microbiome in head and neck cancer containing eleven 16S ribosomal RNA and The Cancer Microbiome Atlas dataset.
- Microbiome from head and neck tissues were able to distinguish tissue types (cancer, cancer-adjacent, non-cancer) using 16S rRNA sequencing and whole genome sequencing datasets.
- Specific bacterial genera correlate with different tumour microenvironment phenotypes.
- High abundance *Fusobacterium* in tumour tissue correlates with better overall survival.

## 1. Introduction

Recent studies have revealed that cancers previously thought to be sterile can contain unique microbial communities. The extent of microbial infiltration varies across different cancer types, with head and neck cancers (HNSC) containing one of the highest level of intratumoral microbial infiltrates while glioblastomas having the least amount of microbes.^1–3^ This “intratumoral microbiota” can refer to bacterial infiltrates found in the extracellular matrix or within the cellular components of the tumour such as cancer, immune and stromal cells.^2^ It is now widely appreciated that intratumoral bacteria can have direct and indirect effects on tumours or the tumour microenvironment (TME).^4–6^ The presence of specific intratumoral bacteria has been reported to influence multiple features of tumour biology including treatment efficacy, local immune composition and activity and promoting tumour metastasis.^7–10^

Direct interaction between specific bacterial species with the tumour and the TME can induce chemoresistance, promote tumour progression, enhance therapeutic responses and modulate anti-tumour immunity through various mechanisms.^11–14^ Bacteria can metabolise an active drug into its inactive form or induce autophagy in cancer cells which can promote chemoresistance.^12–14^ Moreover, specific bacterial species can mount or suppress anti-tumour responses.^15–17^ Most notably, *Fusobacterium nucleatum* colocalises with cancer and immune cells by binding to cell surface receptors such as Toll-like receptor 4 (TLR-4), T-cell immunoreceptor with Ig and ITIM domains (TIGIT) and Carcinoembryonic Antigen-Related Cell Adhesion Molecule 1 (CEACAM-1) receptors, or sugar groups (e.g. tumour expressed Galactose-N-acetylgalactosamine), which may then promote chemoresistance and suppress anti-tumour immunity^13, 15, 18–22^. Alternatively, *Bifidobacterium species* enhance anti-tumour immunity and efficacy of PD-1 immunotherapy responses.^8, 23, 24^

The release of bacterial metabolites such as short chain fatty acids (SCFA), amino acids, vitamins and bile acids can indirectly affect the tumour and the TME.^25, 26^ Butyrate, a SCFA released by anaerobic bacteria through fermentation of carbohydrates, can decrease tumour cell growth and invasion, while increasing CD8^+^ T cell-mediated anti-tumour responses.^27–30^. However, butyrate has also been shown to have pro-tumorigenic effects by inducing senescence-associated inflammatory phenotypes and inhibiting natural killer cell functions.^31, 32^ Bacteria-derived indole and its derivatives (i.e. indole-3-lactic acid) have been shown to suppress anti-tumour immunity by activating immunosuppressive tumour-associated macrophages in treatment-naïve pancreatic cancer, while improving chemotherapeutic and immune-checkpoint inhibitor efficacy in pancreatic cancer and melanoma.^33–35^ Together, these studies demonstrate that the tumour microbiota can influence cancer clinical outcomes in a context-dependent manner.

There are multiple reports describing the microbiota in HNSC.^36–89^ Most of these studies compared the microbiota diversity and bacterial relative abundance between cancer and healthy samples using 16S ribosomal RNA (rRNA) sequencing^36–85, 88, 89^, while two studies additionally correlated the impact of the microbiota with matched transcriptome analysis.^86, 90^ Samples studied include tissues, swabs, and oral fluids (saliva or oral rinse) from cancer and healthy patients. Specifically for HNSC tissue microbiota analysis, samples included cancer, cancer-adjacent (approximately > 5 mm away from the tumour), contralateral, and healthy donor tissue samples.^36–55, 57–60, 85–89^ Most bacteria identified in HNSC are oral commensal bacteria from the genera *Streptococcus, Rothia, Fusobacterium, Haemophilus* and *Prevotella.*^36–38^ However, changes in microbial composition have been identified when cancer samples are compared to healthy controls. In general, there was an enrichment in *Fusobacterium* within cancer tissue samples, that correlated with an inflammatory phenotype.^36, 37, 47^ However, inconsistencies are observed for microbes such as *Streptococcus*, *Actinomyces* and *Prevotella* warranting the need to identify a consensus microbiota signature for HNSC.^37, 38, 43, 54, 85^

In this study, we systematically reviewed the literature and performed a meta-analysis to consolidate the currently heterogenous HNSC-associated microbiota data. Selected 16s rRNA sequencing datasets were analysed consistently to minimise variability between different sample cohorts and adjusted for batch-effects.^91^ These consensus HNSC-associated microbial signatures were then validated using whole genome sequencing (WGS) data from The Cancer Microbiome Atlas (TCMA).^1^ Finally, we correlated the presence of different microbiota signatures with the HNSC tumour microenvironment and clinical outcomes.

## 2. Methods

This study was performed according to the Preferred Reporting Items for Systematic Reviews and Meta-Analyses (PRISMA) Statement.^92^

### 2.1 Search and Study Selection

The following criteria were used to select datasets: 1) Tissue samples, 2) Presence of metadata to distinguish sample types, 3) Illumina short-read amplicon sequencing of 16S rRNA V3 to V5 primers (Figure 1). Database search was performed on 16 August 2022 and datasets after this date were not included (Supplementary Table 1). The risk of biasness assessment was conducted using RoB 2 (β v9) (Supplementary Table 1).

**Figure 1:**
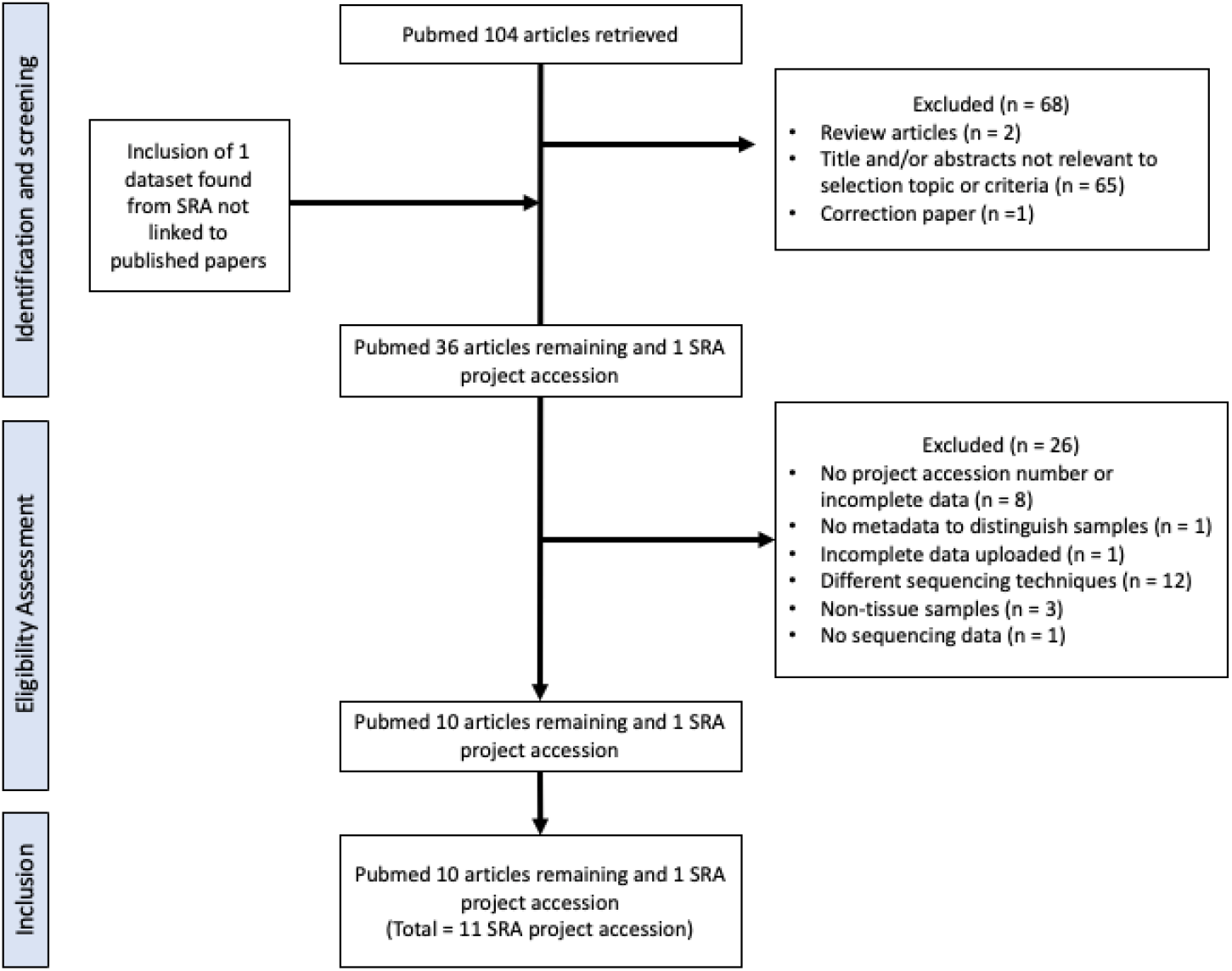
Study selection flow chart.

### 2.2 Download, pre-processing, and analysis of 16S rRNA datasets

Previously published raw sequences were retrieved from the National Center for Biotechnology Information (NCBI) Sequence Read Archive (SRA) using pysradb.^93^ Samples were divided into three main groups – cancer, cancer-adjacent and non-cancer tissues. Cancer tissues are defined as tissues obtained directly from the tumour, while cancer-adjacent tissues are cancer-free regions obtained > 5mm away from cancer tissues. Non-cancer tissues are defined as tissues that were either obtained from healthy patients or contralateral tissues obtained from cancer patients. FASTQ sequences files were obtained from SRA using sratoolkit.^94^ These sequences were processed using QIIME2 DADA2 denoise-paired and reads truncated using the same parameters (trim_left_f = 30, trim_left_r = 30, trunc_q = 15). Sequences from different studies were merged before bacterial Operational Taxonomic Units (OTU) classification using QIIME2 and SILVA reference database (version silva-138-99-nb-classifier).^95^

Raw microbial reads were filtered, central log-ratio (CLR) transformed and batch-adjusted using Phyloseq and MixOmics as described previously.^96–98^ Microbiome datasets are inherently compositional, hence, CLR transformation addresses generates scale-invariant values which allows datasets to remain unaffected by variations in library sizes among samples.^99^ Briefly, low abundance of OTUs were filtered through proportional counts of all samples (< 1%) and minimum counts per sample (< 10). Bacterial OTUs were agglomerated at the genus level before transforming into CLR for their compositional nature.^96, 98^ The CLR-abundance was used for subsequent statistical and discriminant analysis. A total of 903 SRA samples from 11 projects were downloaded (Table 1).

**Table 1:**
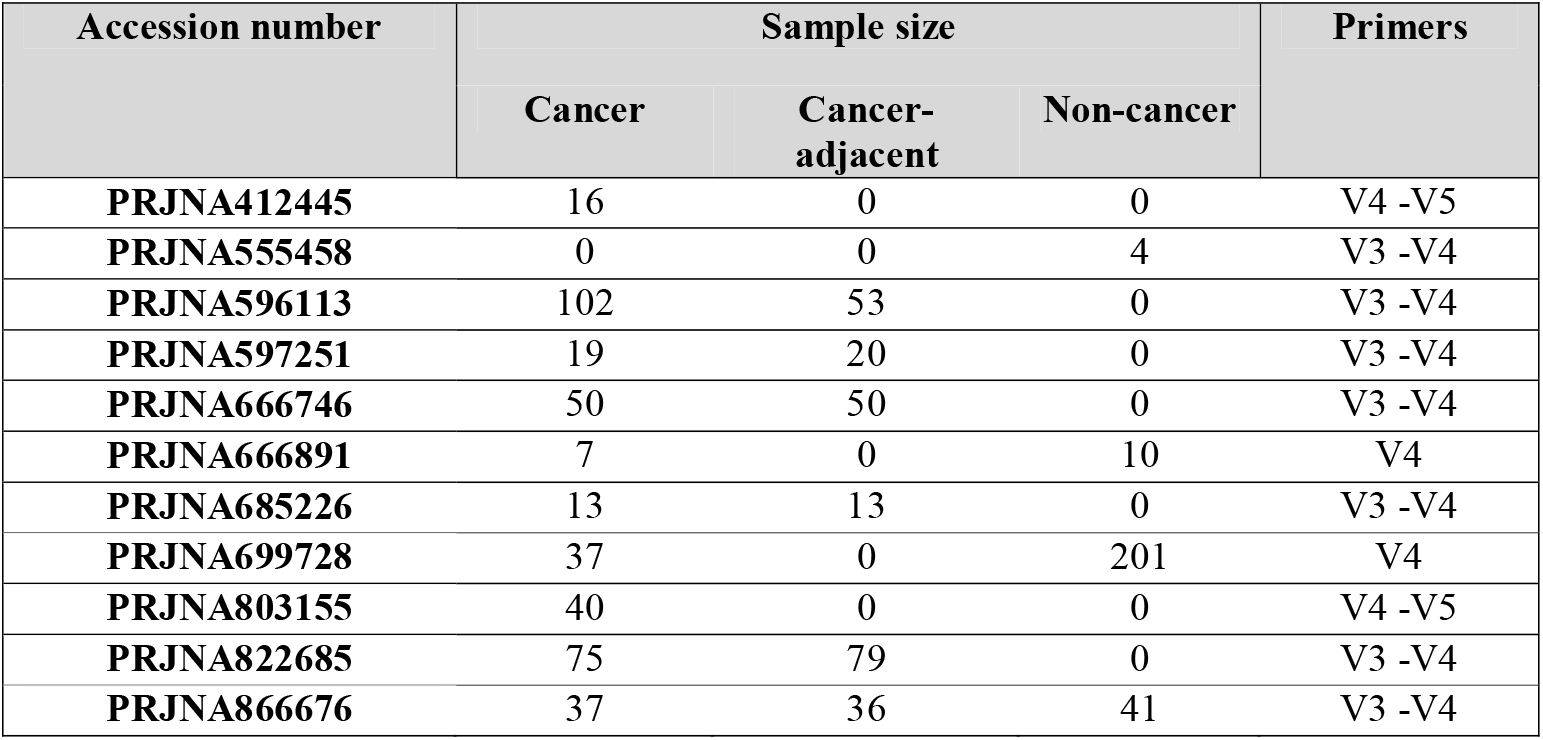
Study accession and sample size post-filtering.

### 2.3 Discriminant analysis of 16S rRNA dataset

To discriminate the microbial signature between sample types, we employed both multivariate and univariate discriminant analysis. For β-diversity analysis, CLR-abundance of all genera were ordinated using Euclidean distance and plotted on a principal component analysis (PCA) using mixOmics R package. β-diversity for each sample were calculated as distance to centroid for each tissue groups using betadisper (vegan v2.6-4). Group and pairwise permutest (vegan v2.6-4, permutations = 9999) was performed to determine if dispersions differed between sample types, while group and pairwise permutational multivariate analysis of variance (PERMANOVA) was performed using adonis2 (vegan v2.6-4, method = “euclidean”, permutation = 9999) and pairwise.adonis2 (pairwiseAdonis, method = “euclidean”, permutation = 9999) to determine statistical differences in β-diversity between groups. Other statistical test such as Analysis of similarities (ANOSIM) (vegan v2.6-4, distance = “euclidean”, permutation = 9999) and Fifty-fifty multivariate analysis of variance (FFMANOVA) (nSim = 9999) were also applied as supplementary to distinguish between sample types.^100, 101^

Multivariate sparse partial linear discriminant analysis (sPLS-DA) was applied on batch-adjusted dataset to identify discriminating genera within each sample type.^96^ The Area Under Curve (AUC) of the Receiver Operating Characteristics (ROC) curve was calculated using mixOmics in Rstudio.^96, 98^ The AUC value served as a quantification of the discriminatory potential between sample types. A higher AUC value, closer to 1, signified a test approaching perfection in its ability to distinguish between the samples. Heatmap of all representative bacteria in each sPLS-DA was presented with sample type clustered according to Euclidean distance and Ward’s linkage.

Univariate Kruskal-Wallis test with Bonferroni multiple comparisons test was also performed to determine microbial genera differences between sample types using microbiomeMarker in RStudio v3.3.0, followed by a post-hoc Wilcoxon test (Mann-Whitney test) with Bonferroni-Dunn multiple comparison test to determine differences between groups (cancer– cancer-adjacent, cancer – non-cancer, non-cancer – cancer-adjacent). Additionally, Wilcoxon matched-pairs signed rank test with Bonferroni-Dunn multiple comparison test was also performed on paired cancer and cancer-adjacent samples.

### 2.4 Functional profiling analysis of 16S rRNA datasets in different sample types

To predict the microbial functions of genera detected from 16S rRNA sequencing between each tissue sample type, Phylogenetic Investigation of Communities by Reconstruction of Unobserved States 2 (PICRUST2) from QIIME2 was applied on raw 16S rRNA reads using MetaCyc database.^102, 103^ Functional abundance was processed and analysed similarly as described for raw microbial reads. Univariate Kruskal-Wallis test and post-hoc Wilcoxon test was performed as previously described to compare differences between groups.

### 2.5 Reanalysis of tissue microbiome data from TCMA

Decontaminated microbial read count derived from The Cancer Genome Atlas (TCGA) HNSC whole genome sequences were obtained from TCMA repository.^1^ Data from a total of 177 cancer (TCGA annotation: primary tumour) and 22 cancer-adjacent (TCGA annotation: solid tumour normal) tissues were obtained from TCMA repository (n = 22 paired cancer and cancer-adjacent samples). Similar to 16S rRNA pre-processing, read counts were agglomerated to the genus before CLR transformation as described in 2.2. As samples were already pre-processed in the TCMA dataset, no further filtering or batch adjustment was required. Microbiome statistical analysis were performed similarly as 16S sequencing datasets. Metadata were obtained from cBioPortal for Cancer Genomics.^104^

### 2.6 Microbiome correlation analysis with tumour microenvironment and survival analysis

The TME immune subtype and 29 functional gene expression signatures (FGES) scores were previously described by Bagaev et al. (2021) using transcriptomics datasets from TCGA.^105^ The 29 FGES represents the major functional components and immune, stromal, and other cellular populations of the tumour.^105^ Pearson’s correlation test was applied to determine the correlation between FGES scores and selected bacteria genera. The four TME immune subtypes were – Desert (D), Fibrotic (F), Immune-enriched (IE), Immune-enriched/Fibrotic (IE/F) (Described in Supplementary Table 2).^105^ Specifically, tissues with IE and IE/F phenotype contains high T-cell infiltration, while D and F phenotypes have low T-cell infiltration (Supplementary Table 2).^105^ Using a cut-off of high (top 35^th^ percentile) and low (bottom 35^th^ percentile) CLR-abundance, the proportion of each patient within the four TME subtypes were determined, and survival analysis was performed. Since there were 153 TCGA-HNSC samples with both FGES/TME subtypes and microbiome datasets, these samples were used for subsequent correlation and survival analysis. Chi-squared (χ^2^) test was performed in Prism9 to determine association between high/low bacterial genera CLR-abundance and proportion of patients within each tumour subtype.

### 2.7 Statistical analysis

For comparisons made between all unpaired tissue groups, Kruskal-Wallis test with Bonferroni’s multiple comparison was used for comparisons made between all tissue groups unless stated otherwise. Post hoc Wilcoxon matched pairs signed rank test with Bonferroni’s multiple comparison was used to compare differences between unpaired tissue samples. For all paired cancer and cancer-adjacent samples, Wilcoxon matched-pairs signed rank test was performed. Univariate and multivariate Cox proportional hazard model was performed using survminer in Rstudio v3.3.0. Statistical analysis was performed using RStudio v3.3.0 and Prism9.

## 3 Results

### 3.1 Multivariate analysis identifies homogenous microbial abundance and functions between cancer and cancer-adjacent samples, contrasting to non-cancer samples

The 16S rRNA amplicon datasets were obtained for 903 head and neck tissue types (396 cancer, 251 cancer-adjacent, and 256 non-cancer) from 11 studies.^37, 38, 41–45, 87–89^ Following sample processing and aggregation of 16S data at the genus level, a total of 177 distinct bacterial genera were identified. Differences in the microbiota and β-diversity between tissue types were assessed using PCA and PERMANOVA test (Figure 2A-2B). The β-diversity index was calculated for cancer (14.6 ± 5.7) and cancer-adjacent (15.0 ± 5.5) tissues, revealing similar levels of β-diversity. In contrast, non-cancer tissues (8.61 ± 5.3) exhibited lower β-diversity (PERMANOVA – Overall R2 = 0.006, p < 0.0001) (Figure 2B). Post-hoc pairwise test identified significant differences in β-diversity between cancer and non-cancer (R^2^ = 0.003, p = 0.002), cancer and cancer-adjacent (R^2^ = 0.005, p < 0.001), and non-cancer and cancer-adjacent samples (R^2^ = 0.007, p < 0.001) (Figure 2B). These findings were consistent with additional multivariate and univariate statistical analysis, ANOSIM (R = 0.027, p = 0.002) and FFMANOVA (p < 0.0001) (Supplementary Table 3).

**Figure 2:**
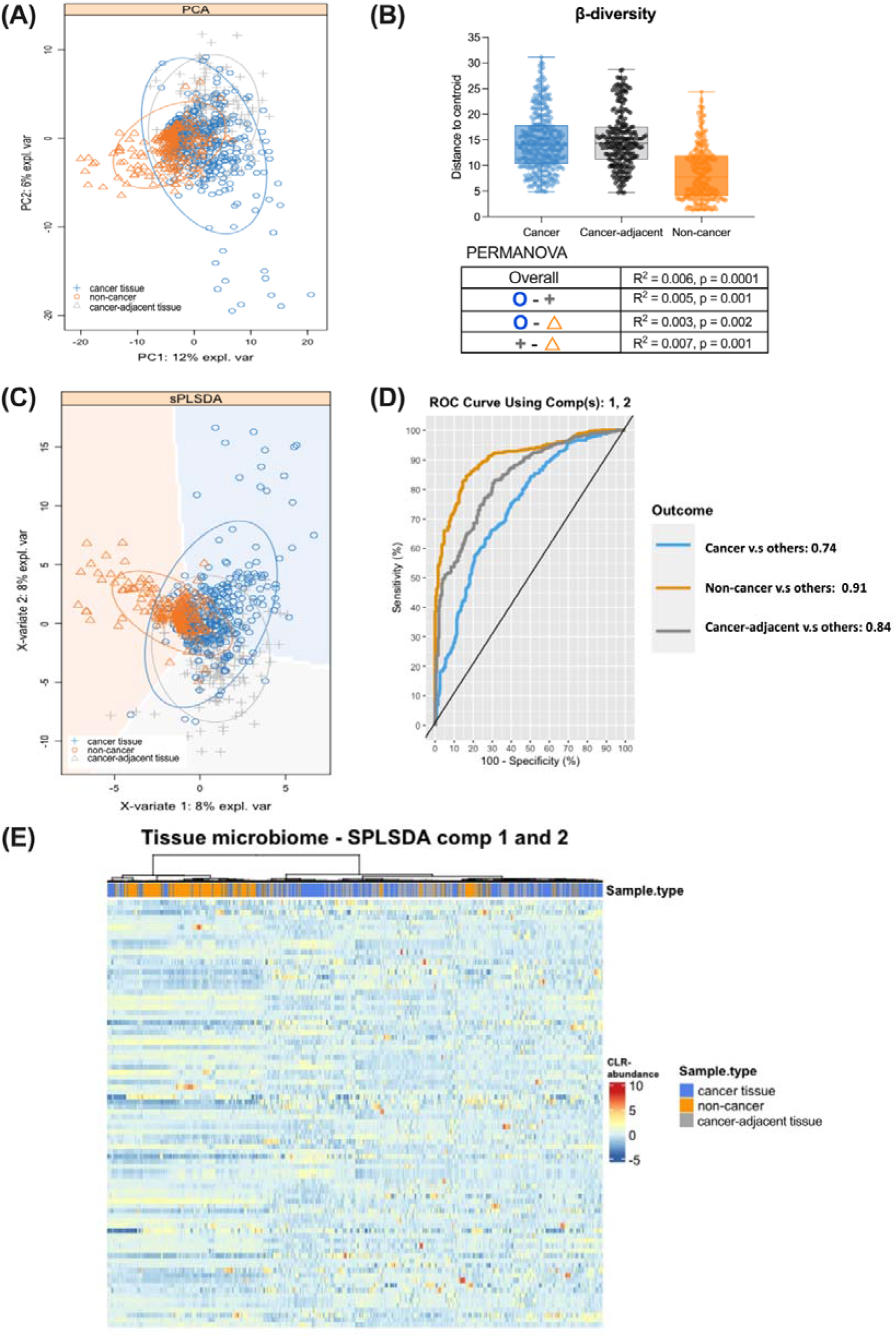
Multivariate discriminant analysis (sPLS-DA and PERMANOVA) of tissue 16S rRNA microbiota to discriminant between cancer, cancer-adjacent and non-cancer tissues. (A) Principal coordinates analysis (PCA) plot of tissue CLR-abundance microbiota based on Euclidean distance. (B) Dispersion of β-diversity (top-right panel) for each sample type, with error bar representing 95% confidence interval. PERMANOVA test was performed with bacterial genera as variable for sample types. (C) sPLS-DA sample plot of 16S rRNA tissue microbiota. Ellipse displays 95% confidence interval for each sample group. The batch-adjusted normalized abundance of tissue microbiota from 16S amplicon sequencing was compared between cancer, cancer-adjacent and non-cancer tissue samples. sPLS-DA identified 116 bacterial genera on component 1 and 2. (D) ROC curve and AUC values determined from sPLS-DA analysis was used to access discriminatory potential of sPLS-DA component 1 and 2. (E) Heatmap representing 86 bacterial genera after sPLS-DA discriminant analysis. Each column and row represent a unique sample and bacterial genera respectively, with OTUs clustered based on Euclidean distance and Ward linkage method.

Multivariate sparse partial least squares discriminant analysis (sPLS-DA) identified 116 representative bacterial genera in sPLS-DA component 1 and 2 which were discriminant between tissue types (Figure 2C-E). The AUC values were computed for different sample comparisons: cancer versus others (AUC = 0.74, p < 0.05), non-cancer versus others (AUC = 0.91, p < 0.05), and cancer-adjacent versus others (AUC = 0.84, p < 0.05). These results demonstrate that sPLS-DA components 1 and 2 (Figure 2D) can effectively differentiate between tissue types. Lastly, majority of cancer and cancer-adjacent samples clustered together and were distinct from non-cancer samples, as determined by Euclidean distance metric (Figure 2E).

### 3.2 Univariate analysis identifies differences in microbial abundance and functions between sample types

Next, unpaired univariate analysis was applied to determine the differences between tissue types. Out of the 177 bacterial genera, 33 were identified as significantly different among tissue types using Kruskal-Wallis test (P_adjust_ < 0.05) (Supplementary Table 4). Notably, 18 of these were also identified as representative bacterial genera in sPLS-DA discriminant analysis (Supplementary Table 4). These 33 genera are denoted as bacterial genera of interest (Supplementary Table 4). The top 20 differentially abundant genera, based on the effect size (η^2^), are presented in Figure 3. Post-hoc unpaired Wilcoxon test with Bonferroni-Dunn’s multiple comparison test was performed on these genera to determine the mean differences in the central log ratio transform (CLR) abundance between tissue types (Figure 3A, Supplementary Table 5). Since most published studies compared cancer to non-cancer, or cancer to cancer-adjacent tissues, we performed post-hoc test for these comparisons (Figure 3B). We identified 27 out of 33 genera as significantly different (P_adjust_ (#) < 0.05) between cancer and non-cancer tissues (Figure 3A-3B, Supplementary Table 5). Non-cancer tissues contained more *Fretibacterium* (CLR-abundance diff. = 1.42, SE = 0.12), *Stenotrophomonas* (CLR-abundance diff. = 0.80, SE = 0.12) and *Tannerella* (CLR-abundance diff. = 0.71, SE = 0.10), while cancer tissue had a greater CLR-abundance of *Neisseria* (CLR-abundance diff. = 2.32, SE = 0.15), *Capnocytophaga* (CLR-abundance diff. = 2.02, SE = 0.15), and *Streptococcus* (CLR-abundance diff. = 1.98, SE = 0.19) (Figure 3A-3B). *Capnocytophaga* abundance in cancer tissues was consistent to previous findings^46, 57, 85^, while contradicting findings were identified for the abundance for *Streptococcus^38, 41, 52, 57, 85^* and *Fusobacterium^38, 41, 42, 52, 57, 58^*.

**Figure 3.**
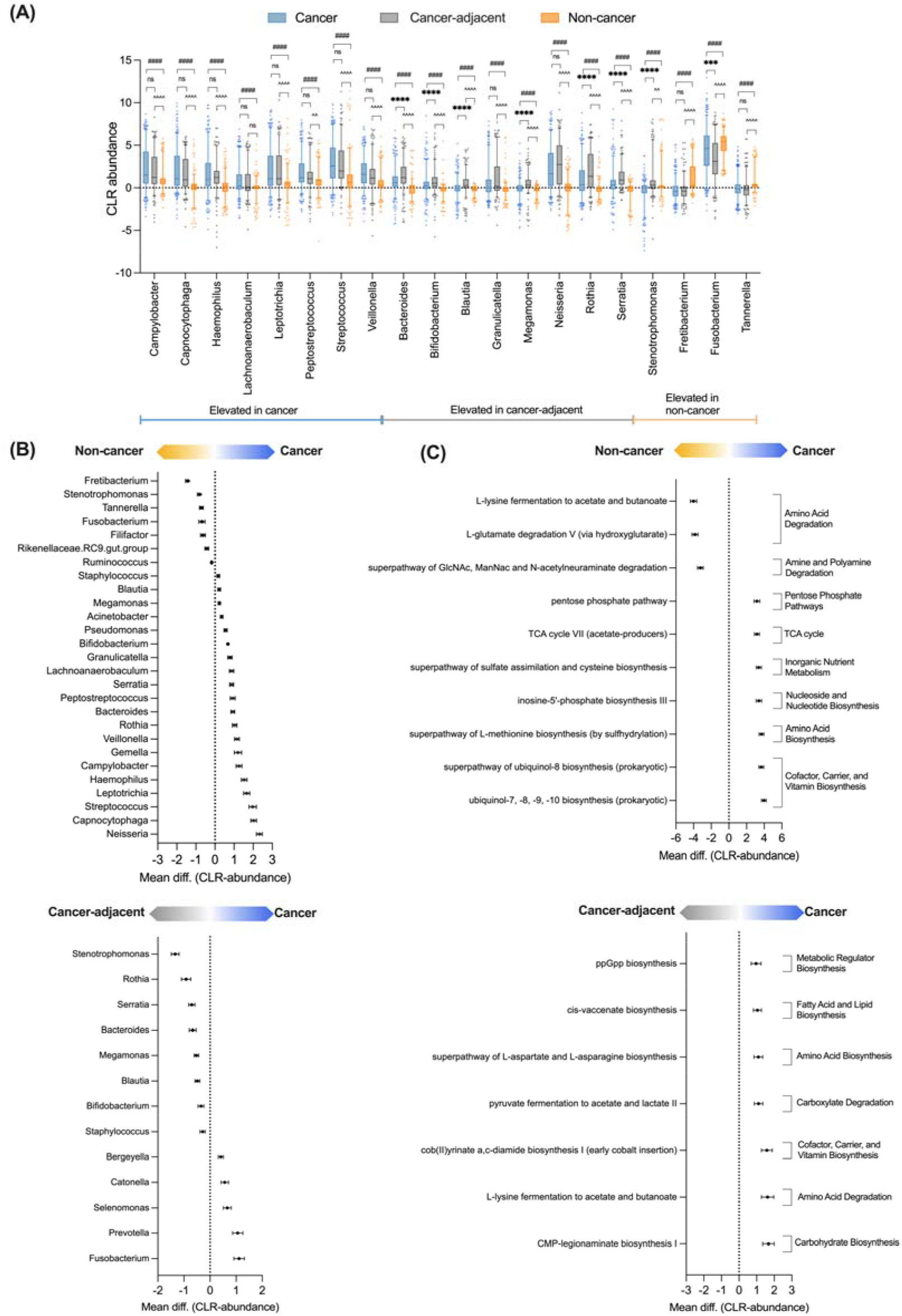
Comparison of bacterial CLR-abundance and functional prediction between sample types. (A) Top 20 bacterial genera (based on effect size) in CLR-normalized abundances between sample groups using Kruskal-Wallis test with Bonferroni’s multiple comparison. 33 out of 177 genera were identified as significantly different (P_adjust_ < 0.05) using Kruskal-Wallis test. Post-hoc Wilcoxon test with Bonferroni-Dunn’s multiple comparison was performed to identify group-wise differences between Cancer – Non-cancer (#), Cancer – Cancer-adjacent (*), Non-cancer – Cancer-adjacent (^). Post-hoc unpaired Wilcoxon test with Bonferroni-Dunn’s multiple comparison for (B) bacterial genera and (C) functional CLR-abundance for Cancer – Non-cancer (Top panel), and Cancer – Cancer-adjacent (Bottom panel).

For cancer and cancer-adjacent tissue, 13 out of 33 bacterial genera were significantly different (post-hoc unpaired Wilcoxon test P_adjust_ (*) < 0.05) (Figure 3A and 3C, Supplementary Table 5). Similar to many studies, *Fusobacterium* (CLR-abundance diff. = 1.11, SE = 0.20) displayed significantly higher CLR-abundance in cancer tissue than cancer-adjacent tissue, while *Rothia* (CLR-abundance diff. = 0.92, SE = 0.18), *Stenotrophomonas* (CLR-abundance diff. = 1.33, SE = 0.15) and *Serratia* (CLR-abundance diff. = 0.70, SE = 0.12) had higher CLR-abundances in cancer-adjacent tissue than cancer tissue (Figure 3A).^36, 37, 43, 45, 52, 55, 58, 59^. Additionally, we found that *Prevotella* was elevated in cancer tissue as compared to cancer-adjacent tissues.^43, 45, 52, 55, 58^ Unlike previous studies, we did not observe any significant differences in *Streptococcus* abundance between cancer and cancer-adjacent tissues.^36, 37, 45, 51, 52, 55, 59^

Lastly, 28 of the 33 top bacterial genera were significantly different (post-hoc unpaired Wilcoxon test P_adjust_ < 0.05) when comparing non-cancer to cancer-adjacent tissue samples (Figure 3A, Supplementary Table 5). Genera *Neisseria* (CLR-abundance diff. = 2.83, SE = 0.19), *Rothia* (CLR-abundance diff. = 1.95, SE = 0.16) and *Streptococcus* (CLR-abundance diff. = 1.95, SE = 0.16) were higher in CLR-abundance in cancer-adjacent, while *Fusobacterium* (CLR-abundance diff. = 1.80, SE = 0.18) and *Prevotella* (CLR-abundance diff. = 1.28, SE = 0.20 were greater in CLR-abundance in non-cancer tissue (Figure 3A).

To provide functional insights to microbial abundance between cancer tissues and other tissue types, we applied Picrust2 to predict possible differences in MetaCyc pathway functional CLR-abundance.^102^ After filtering low abundant functional pathways, we identified a total of 365 MetaCyc pathways. Using Kruskal-Wallis test, 162 MetaCyc pathways were identified as significantly different among sample types (P_adjust_ < 0.05) (Supplementary Table 5). Post-hoc analysis identified 129/162 and 7/162 pathways that were significantly different between cancer – non-cancer, and cancer – cancer-adjacent tissues comparisons respectively (Supplementary Table 6).

Cancer tissues, when compared to non-cancer tissues, were enriched in pathways involving the synthesis of ubiquinol, L-methionine, inosine-5’-phosphate and cysteine and metabolic pathways such as TCA cycle and pentose phosphate pathway, while non-cancer tissues were enriched in the degradation of L-lysine, L-glutamine, N-Acetylglucosamine (GlcNac), N-acetylmannosamine (ManNac), and N-acetylneuraminate (Figure 3C). Cancer tissues were more similar to cancer-adjacent tissues, albeit enrichment was identified in pathways involving biosynthesis of ppGpp (guanosine pentaphosphate and tetraphosphate), cis-vaccenate, L-asparatate, L-asparagine, cob(II)yrinate a,c-diamide and CMP-legionaminate, and enrichment in pathways involving degradation of pyruvate and L-lysine, when compared to cancer-adjacent tissues (Figure 3C).

### 3.3 Paired cancer and cancer-adjacent tissues display similar bacterial abundance differences using multiple sequencing techniques

To understand microbial abundance differences between cancer tissue and cancer-adjacent tissue within the same patients, we performed Wilcoxon matched-pairs signed rank test to identify changes in microbial diversity and abundance within paired tissue samples in the 16S rRNA datasets. Similar to unpaired data analysis, no significant differences in microbial β-diversity was identified between the patient’s paired cancer and cancer-adjacent tissues (Supplementary Figure 2).

However, 76 bacterial genera were significantly different between paired tissue samples (Figure 4A, Supplementary Table 7). Bacterial genera with the greatest differences in CLR-abundance were then identified by using a cut-off of > 0.4 and < -0.4 (Figure 3A). Using this cut-off, we found that *Fusobacterium, Prevotella, Alloprevotella, Catonella, Selenomonas and Treponema* were elevated in cancer tissue vs cancer-adjacent tissue, while *Stenotrophomonas, Rothia, Granulicatella, Serratia, Anoxybacillus, Actinomyces and Bacteroides* were greater in cancer-adjacent tissue compared to cancer tissue (Figure 4A-4B). Similarly, nine of these bacteria were also found to be significantly different in unpaired tissue analysis (Supplementary Table 4 and 7). Contrary to published studies on unpaired samples, *Streptococcus,* an abundant oral commensal, was not significantly different in our paired sample analysis.^36, 37, 45, 51, 52, 55, 59^

**Figure 4:**
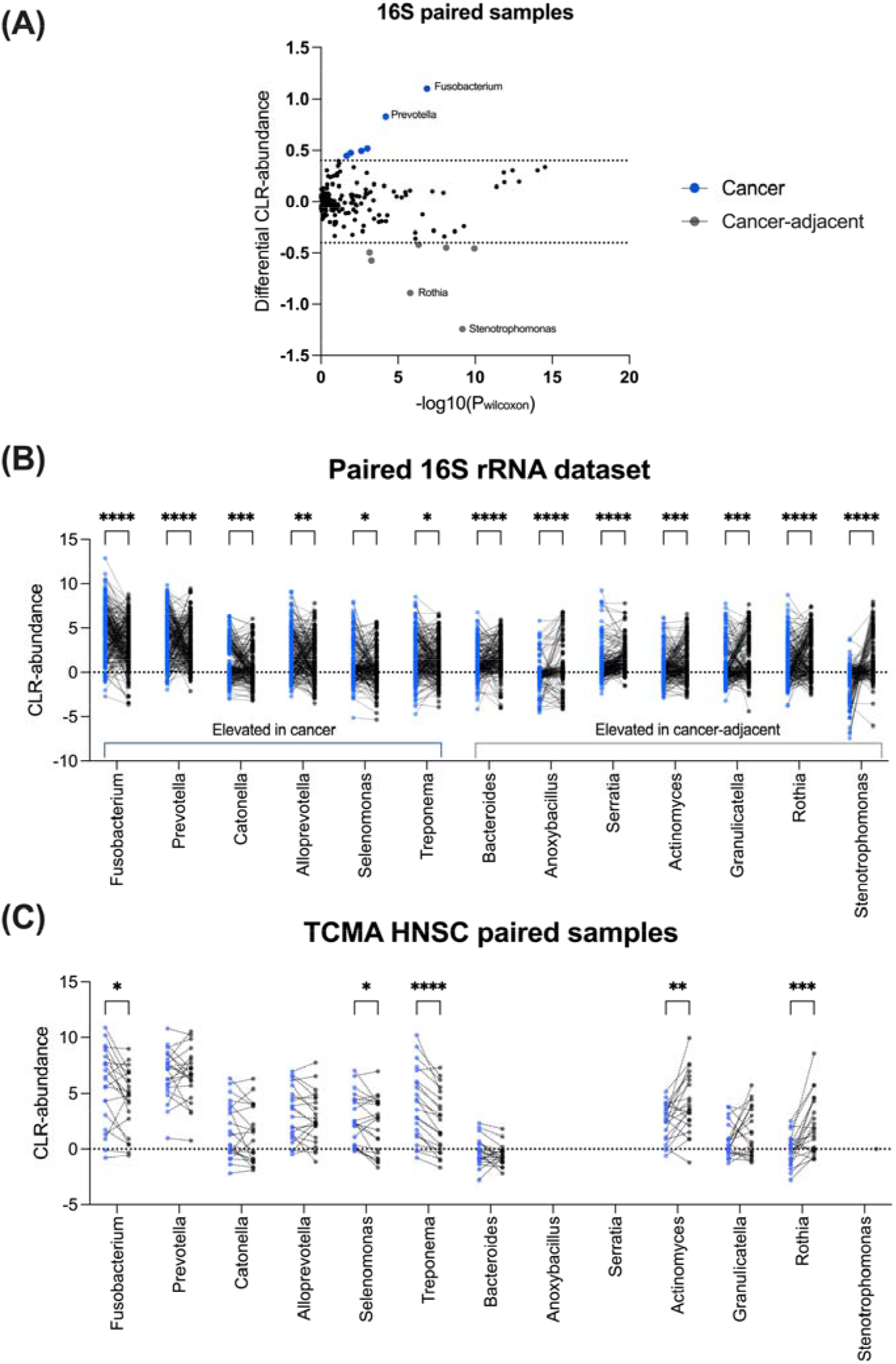
Comparison of tissue microbiota in paired cancer and cancer-adjacent tissue samples using different sequencing datasets. **(A)** Paired Wilcoxon matched-pairs signed rank test on paired 16S rRNA sequencing cancer and cancer-adjacent tissue samples. 76 bacteria were significantly different in sample groups (p < 0.05) using paired Wilcoxon matched-pairs signed rank test and 13 bacteria genera were identified as top bacteria with differential CLR-abundance (Diff. CLR-abundance > 0.4 or < -0.4). Blue and red dot points represent bacteria that were higher in abundance in cancer and cancer-adjacent tissues respectively. CLR-abundance of paired cancer and cancer-adjacent samples from **(B)** 16s rRNA sequencing and **(C)** TCMA WGS sequencing datasets. Wilcoxon matched pairs signed rank test was performed for both 16s rRNA (n = 287) and TCMA (n = 22) datasets. *p < 0.05, **p < 0.01, ***p < 0.001, ****p < 0.0001.

To validate this finding, we probed the publicly available TCMA dataset, a repository containing microbiota reads derived from WGS of tissue samples.^1^ Similar to the 16S rRNA dataset, we observed that cancer tissues from TCMA displayed significantly (p < 0.05) higher CLR-abundance for genera *Fusobacterium, Selenomonas* and *Treponema*, while *Rothia* and *Actinomyces* were elevated (p < 0.05) in cancer-adjacent tissues (Figure 4D). In the TCMA dataset, *Anoxybacillus, Serratia,* and *Stenotrophomonas* were not present due to pre-analysis filtering, while no significant differences in CLR-abundance were observed for *Prevotella*, *Catonella*, *Alloprevotella*, and *Bacteroides* (Figure 4C). Notably, similar trend in CLR-abundance between cancer and cancer-adjacent samples was still observed for *Prevotella*, *Catonella*, and *Alloprevotella* in TCMA dataset. Overall, 16S rRNA and TCMA WGS dataset showed similar trend for most bacteria genera, regardless of sequencing techniques.

### 3.4 Tissue microbiota diversity correlates with cancer functional gene expression signatures

Since *Fusobacterium, Selenomonas, Treponema, Actinomyces,* and *Rothia* displayed significant differences between paired cancer and cancer-adjacent tissues, we performed correlation analyses to investigate the possible relationship between these genera and the tumour transcriptional profile and patient clinical features found in matched TCGA patients (n = 156).^105^ Here, TCGA transcriptomic data were classified into 29 functional gene expression signatures (FGES), which represent major functional components and characteristics of cancer cell populations.^105^ These 29 FGES can then be used to further classify cancers into four major immune subtypes (Desert, Fibrotic, Immune-enriched/non-fibrotic, and Immune-enriched/fibrotic).^105^ We correlated the CLR-abundance of *Fusobacterium, Selenomonas, Treponema, Actinomyces,* and *Rothia* from TCGA-HNSC patients with their respective FGES scores and immune subtype.

We first correlated CLR-abundance with the FGE signatures. The CLR-abundance of *Fusobacterium* correlated (r > 0.3, p < 0.0001) with FGES related to angiogenesis, neutrophils and granulocyte traffic (Figure 5A). Other FGES such as matrix remodelling, protumour cytokines, MDSC traffic, M1 signature, antitumour cytokine, MHCI and EMT signatures also positively correlated (p < 0.05) to CLR-abundance of *Fusobacterium* (Figure 5A). The CLR-abundance of *Selenomonas* showed a positive correlation (p < 0.05) to angiogenesis, neutrophil signature, granulocyte traffic and antitumour cytokines signatures, while negatively correlating (p < 0.05) to B cells (Figure 4A). Lastly, CLR-abundance of Treponema displayed a negative correlation (p < 0.05) to endothelium, T reg traffic, T reg, MHCII, Coactivation molecules, B cells, NK cells, Effector cells and T cells, while positively correlating to (p < 0.05) neutrophils and granulocyte traffic (Figure 5A).

**Figure 5:**
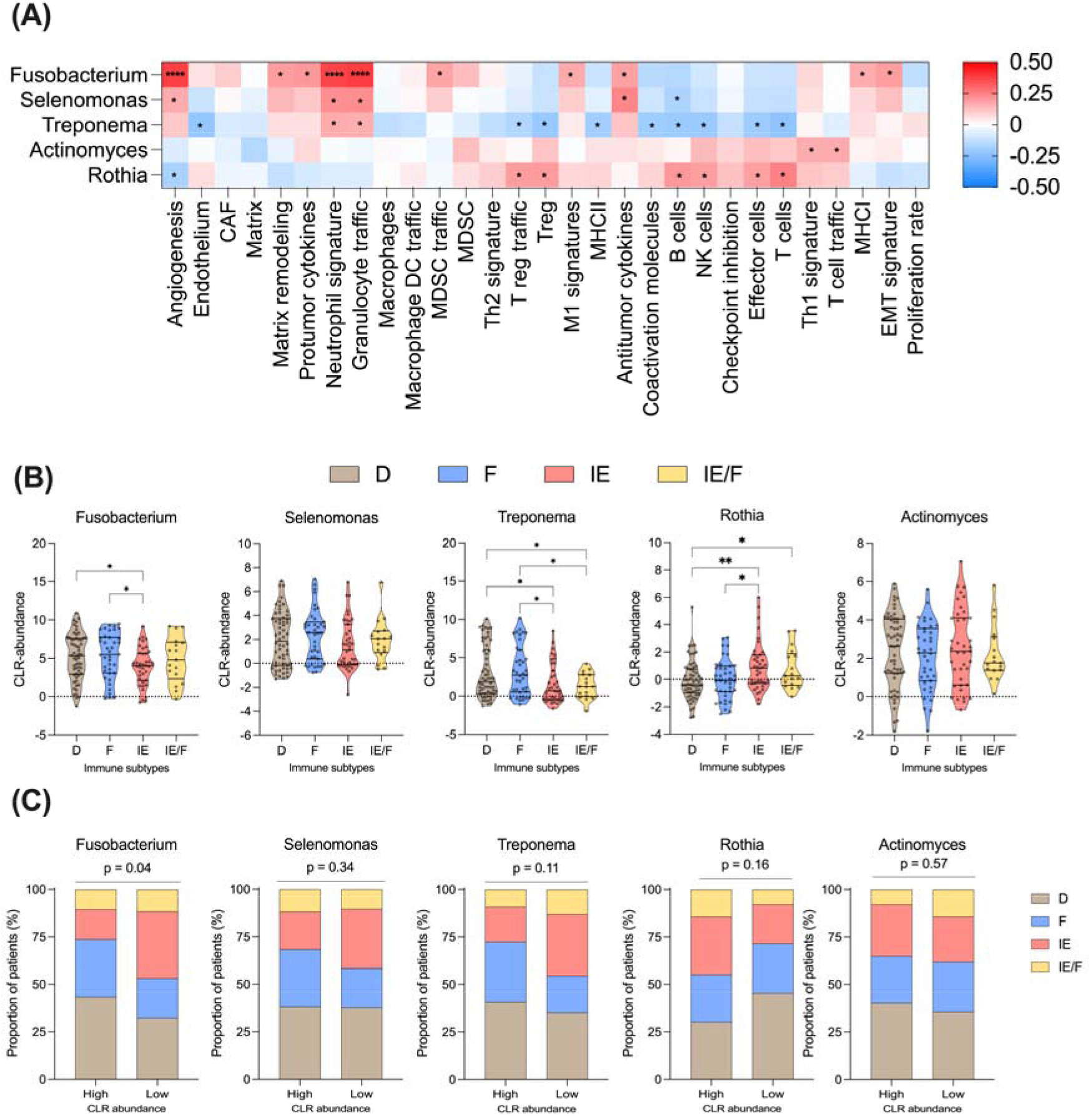
Correlation analysis of *Fusobacterium, Selenomonas, Treponema, Rothia* and *Actinomyces* to the tumour transcriptional profiles. (A) 29 functional gene expression (FGES) signature scores derived from Bagaev et al (2021) were used to correlated with CLR-abundance of genera *Fusobacterium, Selenomonas, Treponema, Actinomyces, and Rothia,* using Pearson’s correlation method. Asterisk (*) represents significant correlation (p < 0.05), and red and blue scales represents positive and negative correlation respectively. **(B)** The CLR-abundance of each bacterial genera within each tumour microenvironment immune subtype (D – Desert, F – Fibrotic, IE – Immune-enriched/Non-fibrotic, IE/F – Immune-enriched/Fibrotic). Kruskal-Wallis test with uncorrected Dunn’s test was performed to compare CLR-abundance in all immune groups. *p < 0.05, **p < 0.01. **(C)** The proportion of patients in each tumour immune subtype with high and low CLR-abundance in each bacterial genera. High and low bacteria CLR-abundance groups were determined by upper and lower 35% quartiles respectively. Chi-squared test was performed to determine association between high/low bacterial genera CLR-abundance and proportion of patients in each tumour subtype.

Next, we investigated how CLR-abundance correlated to tissue immune subtyping. Cancer tissues classified as immune deserts (D) and fibrotic (F) which lack immune cell enrichment correlated with higher *Fusobacterium* and *Treponema* CLR-abundance. On the other hand, cancer tissues that are immune-enriched / non-fibrotic (IE) or immune-enriched / fibrotic (IE/F) correlated with greater *Rothia*. No significant correlation in immune subtypes were observed for *Selenomonas* and *Actinomyces* (Figure 5B). To identify the differences in immune subtypes between high and low CLR-abundance of each bacterial genera, we further segregated patients based on the upper and lower 35% CLR-abundance quartiles. As expected, patients with IE and IE/F tumour subtypes showed significant association with low CLR-abundance of *Fusobacterium* (chi-square test, p = 0.04). While not reaching statistical significance, more patients with IE and IE/F tumour subtypes have low CLR-abundance of *Selenomonas* (chi-square test, p = 0.33) and *Treponema* (chi-square test, p = 0.11), opposite to high CLR-abundance for *Rothia* (Figure 5C). Conversely, patients with D and F subtypes had higher CLR-abundance of *Fusobacterium*, *Selenomonas* or *Treponema* (Figure 5C). Lastly, the proportion of patients in each immune subtype were similar in high and low CLR-abundance *Actinomyces* groups. Taken together, these show that *Fusobacterium*, *Selenomonas* or *Treponema* are associated with poor T-cell infiltration compared to *Rothia* which may have implications in selecting patients suitable for immunotherapy.

### 3.5 Evaluation of microbiota abundance with clinical features and survival

Univariate and multivariate Cox proportional hazard models were used to investigate the association between the intratumoral microbiota and clinical features. Univariate Cox proportional hazard model identified that current smokers (HR 2.235, 95% CI 1.146 – 4.359, p = 0.018), HPV-negative (HR 2.273, 95% CI 1.158 – 4.459, p = 0.017), and low CLR-abundance of *Fusobacterium* (HR 0.8883, 95% CI 0.8183 – 0.9642, p = 0.005) were risk factors for reduced overall survival (Table 2). Further multivariate Cox proportional hazard models identified that HPV-negative (HR 2.853, 95% CI 1.1991 – 6.7882, p = 0.0178) and low CLR-abundance of *Fusobacterium* (Continuous: HR 0.8482, 95% CI 0.7758– 0.9273, p = 0.0003; Low: HR 2.579, 95% CI 1.3687 – 4.860, p = 0.0034) were independent hazards for overall survival, but not current smokers (Table 2).

**Table 2:**
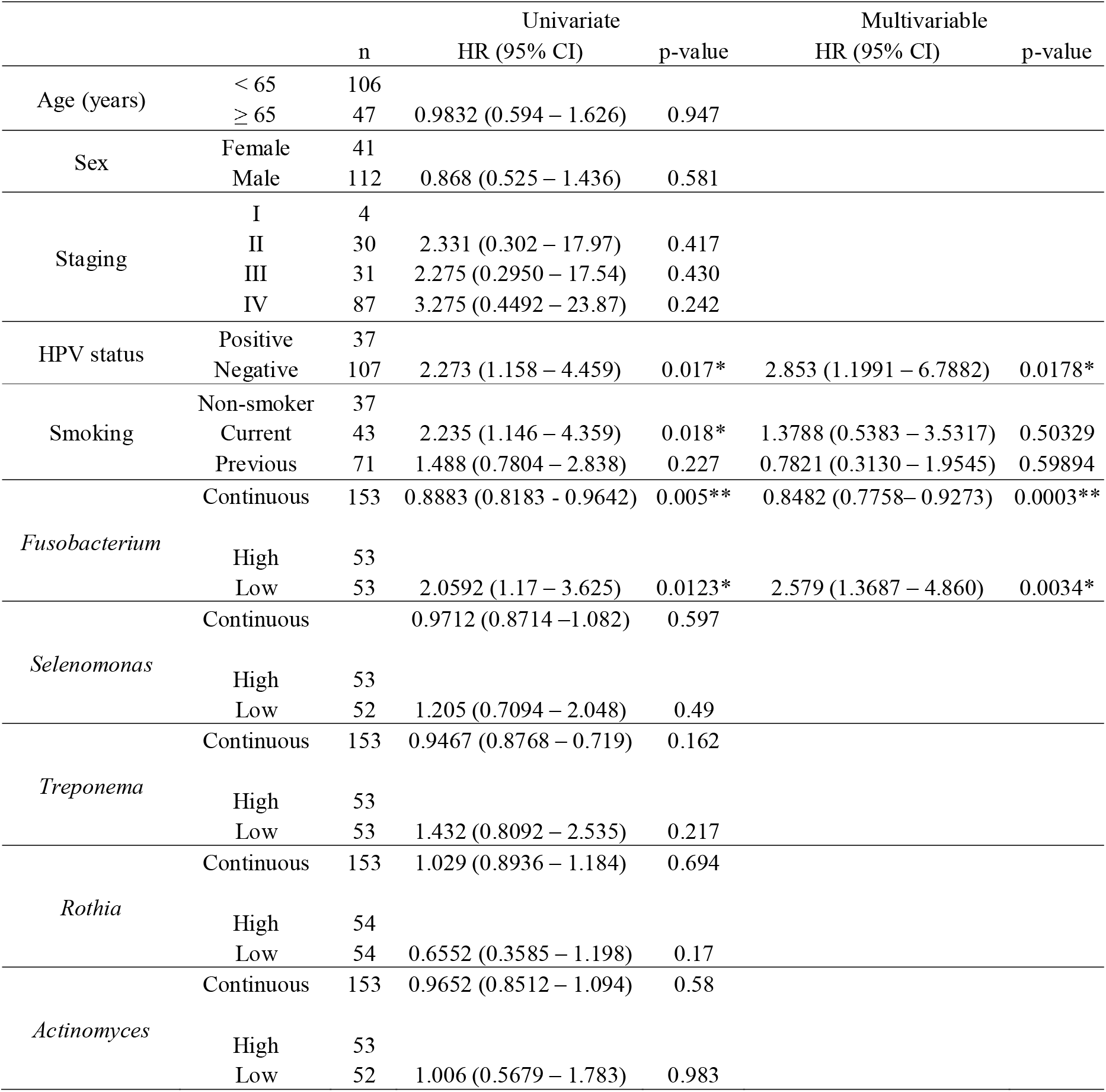
Univariate and multivariable Cox proportional hazard models for overall survival.

## 4 Discussion

Several studies have investigated the microbial signature in HNSC using different sequencing approaches and sample types, such as tissues, swabs, and oral fluids. However, these studies have reported inconsistent findings regarding the presence of specific bacterial genera. Consequently, a consensus microbial signature for head and neck tissues has yet to be established. In this study, we aimed to address this gap by conducting a meta-analysis of 11 studies and presenting a consensus tissue microbiota signature for head and neck tissues. We analyzed 16S rRNA sequencing datasets from 903 tissue samples, including 396 cancer tissues, 251 cancer-adjacent tissues, and 256 non-cancer tissues. Our analysis revealed significant differences in the abundance of 33 bacterial genera among the various tissue types. Specifically, we observed that cancer tissues and cancer-adjacent tissues exhibited greater similarity to each other compared to non-cancer tissues. These findings suggest distinct microbial profiles in cancer and cancer-adjacent tissues compared to non-cancer tissues. Non-cancer tissues exhibited the lowest differences in β-diversity and contained elevated levels of bacterial genera such as *Tannerella*, *Fretibacterium*, *Stenotrophomonas*, *Fusobacterium*, and *Prevotella* (Figure 6A). While cancer and cancer-adjacent tissues displayed similar microbiota based on β-diversity indexes, further analysis using paired and unpaired univariate methods enabled differentiation of these tissues at the genera level (Figure 6B). Importantly, these abundance signatures were validated using additional data from TCMA. Matching TCMA samples with transcriptomic data derived from TCGA) and clinical features provided insights into the contributions of individual genera in HNSC. Notably, we found that a high abundance of *Fusobacterium* was associated with better overall survival in HNSC patients Overall, our study contributes to the establishment of a consensus tissue microbiota signature for HNSC, shedding light on the distinct microbial profiles in different tissue types and their potential implications for clinical outcomes.

**Figure 6:**
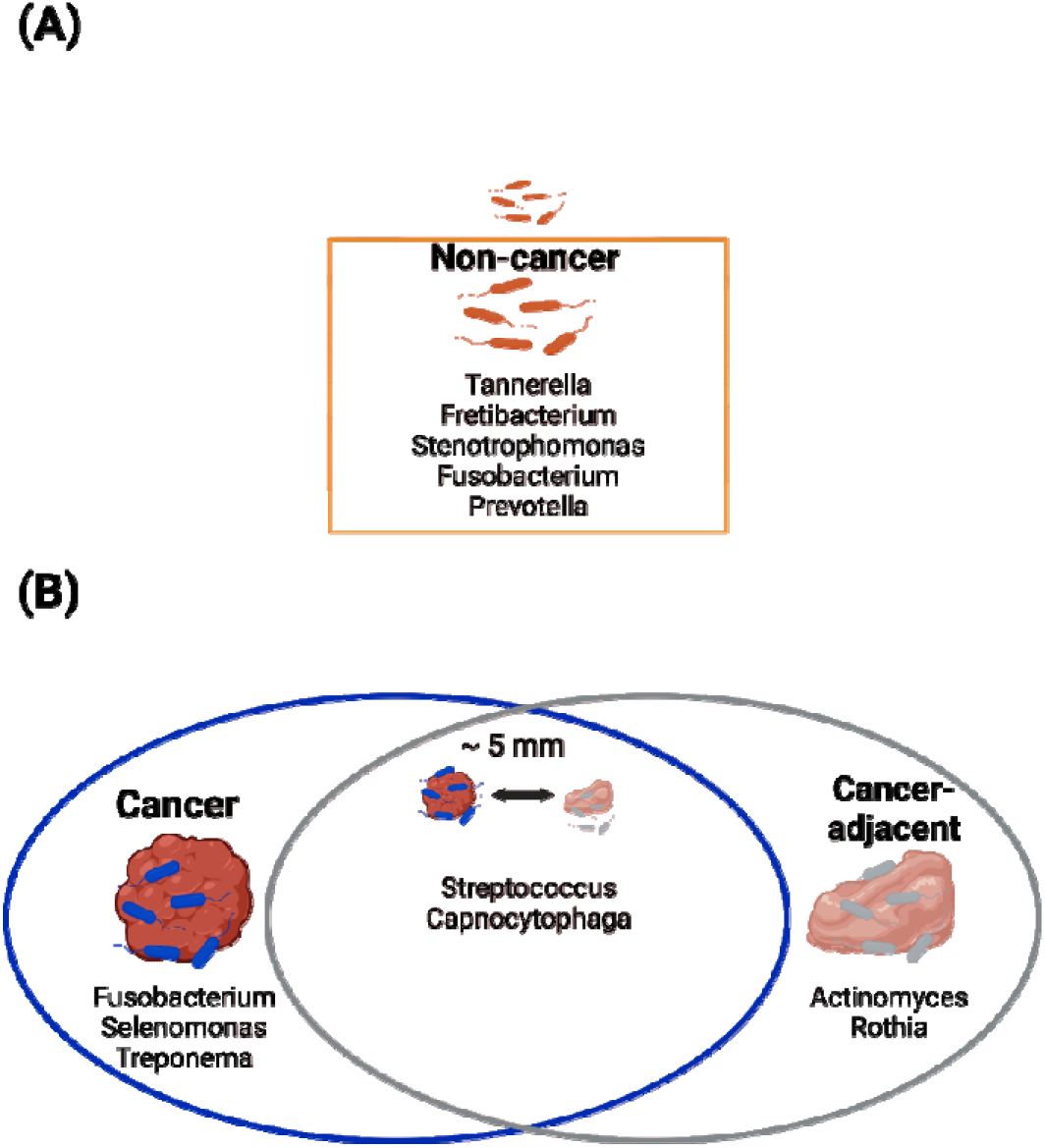
Summary of bacteria genera within cancer, cancer-adjacent and non-cancer tissue samples. **(A)** Elevated microbiota within non-cancer tissues compared to cancer and cancer-adjacent tissues. **(B)** Elevated bacteria genera between cancer and cancer-adjacent tissues.

Both multivariate and univariate discriminant analyses was able to differentiate different tissue sample types based on microbial abundance. As previously reported, cancer and cancer-adjacent tissues were more similar in microbial diversity when compared to non-cancer tissues.^36–38, 42, 43, 45, 49, 51, 52, 55, 57–59, 85^ At the genus level, both paired and unpaired abundance analysis of cancer and cancer-adjacent tissues showed consistent enrichment for *Fusobacterium* and *Rothia* in cancer tissues.^36, 37, 45, 52, 55, 58, 59^ In contrast, *Prevotella* was enriched within cancer tissues compared to cancer-adjacent tissues, and no differences were observed for Streptococcus.^36–38, 41, 43, 45, 51, 52, 55, 57–59, 85^

Previous studies have reported conflicting result where *Fusobacterium* was more in cancer tissues as compared to non-cancer and cancer-adjacent tissues.^38, 42, 52, 57, 58^ However, we found that Fusobacterium was most abundant in non-cancer tissues. *Fusobacterium* is an abundant commensal bacteria found largely in the oral cavity (buccal, hard palate, gingiva, tonsils, tongue) and saliva of healthy individuals, suggesting a potential role within the healthy oral microbiota.^106–108^ *In vitro* experiments in HNSC cell lines showed that *Fusobacterium nucleatum* infection promotes cancer cell invasion, proliferation, autophagy, and PD-L1 expression.^109–113^ It is unknown whether there are strain and species level differences found in *Fusobacterium* isolated in cancer and non-cancer tissues to explain such seemingly contradictory findings. Additionally, non-cancer tissue from cancer patients may also have different tissue microbiota profiles from healthy donor tissues which is currently unavailable for this study. Also, most of the experiments showing an oncogenic role for *F. nucleatum* were carried out *in vitro* and thus did not consider a potential mitigating role of the immune system. Moreover, the abundance of *F. nucleatum* both in absolute terms and relative to other bacteria present in the tumour microbiota might influence the oncogenic potential of *F. nucleatum*. Further experiments are required to evaluate the role of *Fusobacterium* in HNSC.

We observed that *Streptococcus*, another highly abundant oral commensal genera^106–108^, was increased specifically in cancer and cancer-adjacent tissue when compared to non-cancer tissues. However, there was no significant difference in *Streptococcus* abundance between cancer and cancer-adjacent tissues. Within the oral cavity, certain pathogenic *Streptococcus* species, like *S. mutans*, can contribute to periodontitis by acidifying the environment.^114^ In oral cancer, *S. mutans* has been shown to promote tumour proliferation and invasion, potentially through upregulation of IL-6 in infected cells^115^. On the other hand, *Streptococcus* species from the *mitis* (*S. oralis, S parasanguinis, S.mitis*) and *sanguinis* (*S. sanguinis, S. gordonii*) groups, can break down lactic acid or pyruvate into hydrogen peroxide, thereby antagonising pathogenic species such as *S. mutans*.^114^ In oral cancer, *S. mitis, S. salivarius, S. anginosus* were found to display anti-tumour effects, including reducing cancer cell viability and promoting CD8^+^ cytotoxic T cell responses^116–120^. These findings indicate that the abundance of specific *Streptococcus* species may contribute to pathogenesis, disease severity, or exert anti-tumour effects. It is important to note that these studies underscore the limitations of identifying microbiota at the genus level using short-read 16S rRNA sequencing. To address these limitations, recent advances in sequencing technologies such as long-read 16S rRNA amplicon sequencing (e.g., PacBio, Nanopore) or shotgun metagenomics can be employed to reveal species- or strain-specific diversity within the microbiota.^121–123^ Such advancements can provide a more comprehensive understanding of the specific species and strains that play a role in oral cancer pathogenesis and anti-tumour effects.

To compare the metabolic potential of different head and neck tissue types, a functional prediction analysis was performed using PICRUSt2 on the 16S rRNA sequencing data. The analysis revealed an enrichment of several amino acids and metabolites, including L-aspartate, L-asparagine, acetate, butanoate, and lactate, in cancer tissues compared to non-cancer and cancer-adjacent tissues. L-aspartate and L-asparagine, as substrates for nucleotide biosynthesis and regulators of amino acid homeostasis and anabolic metabolism, have been reported to promote tumour proliferation.^124–126^ Butanoate, acetate, and lactate can serve as energy sources for cells by converting into acetyl-CoA, which can then be utilized in the tricarboxylic acid (TCA) cycle to produce ATP.^127–130^ The role of butanoate in tumorigenesis depends on the specific tumour and the TME, as it can exhibit tumour-promoting or suppressive properties. ^31, 131–133^ Lactate, a well-studied metabolite produced by both cancer cells and bacteria, can modulate the TME by inactivating natural killer cells, promoting polarisation of M2-like tumour-associated macrophages, and stimulating the growth of T-regulatory cells.^134^ Collectively, these findings suggest that bacteria infiltrating HNSC tissues possess functional capacities that may promote cancer progression. Further validation studies are warranted to better understand the role of these metabolic pathways in HNSC and the contribution of bacteria in shaping the TME.

We further explored the relationship between the abundance of the five cancer-associated bacterial genera, *Fusobacterium, Selenomonas, Treponema, Actinomyces,* and *Rothia*, and the TME phenotype and clinical outcomes. *Fusobacterium* was associated with a lack of T-cell immune infiltration in HNSC, similar to colorectal and oesophageal cancers.^21, 135–137^ Furthermore, *Fusobacterium* can chemoattract neutrophils via release of SCFA and can also modulate neutrophils and endothelial cell functions *in vitro*.^138–142^ Interestingly, we observed that patients with low levels of *Fusobacterium* within the tumour tissue had shorter overall survival, consistent to previous reports in HNSC.^45, 60, 143^ In contrast, opposite findings have been reported for colorectal, gastric and oesophageal cancers, suggesting that *Fusobacterium* may have a different role in HNSC.^144–147^ We also found that *Treponema* correlated with an lack of immune infiltration in HNSC. Although the effect of *Treponema* infiltration in HNSC is still unknown, these bacteria have been associated with an upregulation of immune suppressive cells and can suppress innate immune responses.^148–150^ In our analysis, *Rothia* was found to correlate with an immune-enriched TME. Limited information is available regarding the role of *Rothia* in cancer; however, *Rothia dentocariosa* has been shown to induce Toll-like receptor 2 (TLR-2) mediated TNF-alpha inflammatory response in human embryonic kidney cells and THP-1 monocytes.^151^ *Selenomonas* and *Actinomyces* did not significantly correlate with TME subtypes in our analysis. However, *Selenomonas sputigena* infected gingival epithelial cells can promote neutrophil and monocyte recruitment.^152^ *Actinomyces* has been associated with young-onset colorectal cancers, showing a preferential localisation with cancer-associated fibroblasts in the TME.^153^ These findings underscore the importance of validating and understanding the underlying mechanisms through which these bacteria can modulate the tumour microenvironment in HNSC.

This study represents the first comprehensive comparison of 16S rRNA (V3-V5) microbial sequencing across multiple studies to identify consensus HNSC-associated microbiota signatures in cancer, cancer-adjacent and non-cancer tissues. To ensure consistency, a uniform bioinformatics approach was employed. However, it is important to acknowledge the inherent limitations of this study. Variations in sample collection, preparation, and sequencing among different laboratories introduce batch effects that could contribute to the inconsistencies observed across different reports. To mitigate these effects, we applied PLSDA-batch adjustment to the pooled datasets.^91^ Conventional short-read 16S rRNA sequencing provides information only up to the genus level, which restricts the ability to identify specific bacterial species or strains that may be relevant to disease outcomes.^121^ Overcoming this limitation would require advanced sequencing technologies such as long-read 16S rRNA amplicon sequencing or shotgun metagenomics to reveal species- or strain-level diversity within the microbiota. Furthermore, the availability of complete clinical metadata in published datasets reporting 16S rRNA sequencing is limited, restricting our ability to make comprehensive clinical associations. Therefore, our clinical associations were primarily based on TCMA/TCGA datasets. Despite these limitations, this study confirms distinct differences in the microbiota composition among cancer, cancer-adjacent and non-cancer HNSC tissue samples. The strength of our study lies in the meta-analysis of a substantial number of samples, totalling 903. Additionally, our analysis indicates that a high load of *Fusobacterium* within HNSC tissues may be associated with a favourable survival outcome. The correlation analysis of the microbiota with functional predictions, functional gene enrichment signature, and immune subtyping of the tumour and TME provides novel avenues for further exploration.

In conclusion, our study establishes a consensus microbial signature for head and neck tissues, shedding light on the distinct microbial profiles present in head and neck cancer (HNSC). These findings have the potential to serve as targets for future treatment approaches in HNSC. Nevertheless, it is crucial to acknowledge the limitations identified in our study and recognize the need for further research to address these limitations. Additional investigations are required to gain a deeper understanding of the functional implications of the identified microbiota differences in HNSC. By addressing these gaps, we can advance our knowledge and pave the way for more effective therapeutic interventions in HNSC.

## 6. Author Contributions

Conceptualisation K.Y.,K.F.; methodology, investigation, and data analysis, K.Y.,R.L.,F.W., E.S., G.B., and L.M.; resources A.P.,P.W. and S.V.; writing - original draft preparation, K.Y., E.S., S.V., and K.F; writing-review and editing, K.Y., G.B., E.S., A.P., P.W., R.V., S.V., and K.F.; supervision R.V.,A.P., S.V., and K.F.; funding acquisition, A.P.,P.W., and S.V.; All authors have read and agreed to the published version of the manuscript.

## 7. Acknowledgments

This work is supported by an NHMRC investigator grant APP1196832 to P.W., a The Garnett Passe and Rodney Williams Senior Fellowship to S.V., and The University of Adelaide Postgraduate Research Scholarship to K.Y., R.L., F.W and L.M. Illustration in Figure 6 was generated using Biorender.

## 8. Data Availability Statement

The data used to support the findings of this study are included within the article and within supplementary material.

## Supplementary Figures

**Figure S1:**
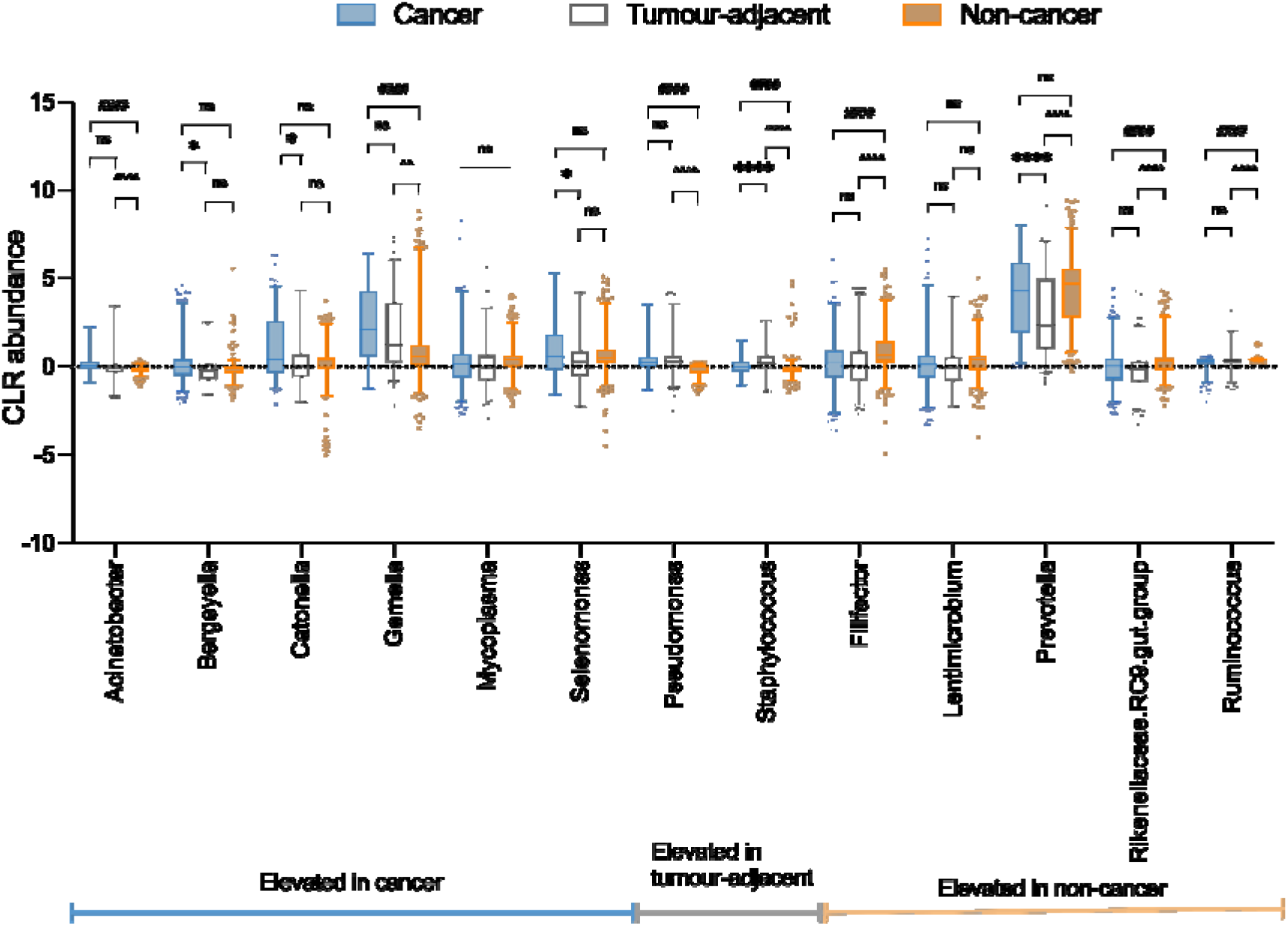
CLR-normalized abundances for remaining 13 bacteria between sample groups using unpaired Kruskal-Wallis test with Bonferroni’s multiple comparison. Post-hoc Wilcoxon test with Bonferroni-Dunn’s multiple comparison was performed to identify group-wise differences between Cancer – Non-cancer (#), Cancer – Cancer-adjacent (*), Non-cancer – Cancer-adjacent (^).

**Figure S2:**
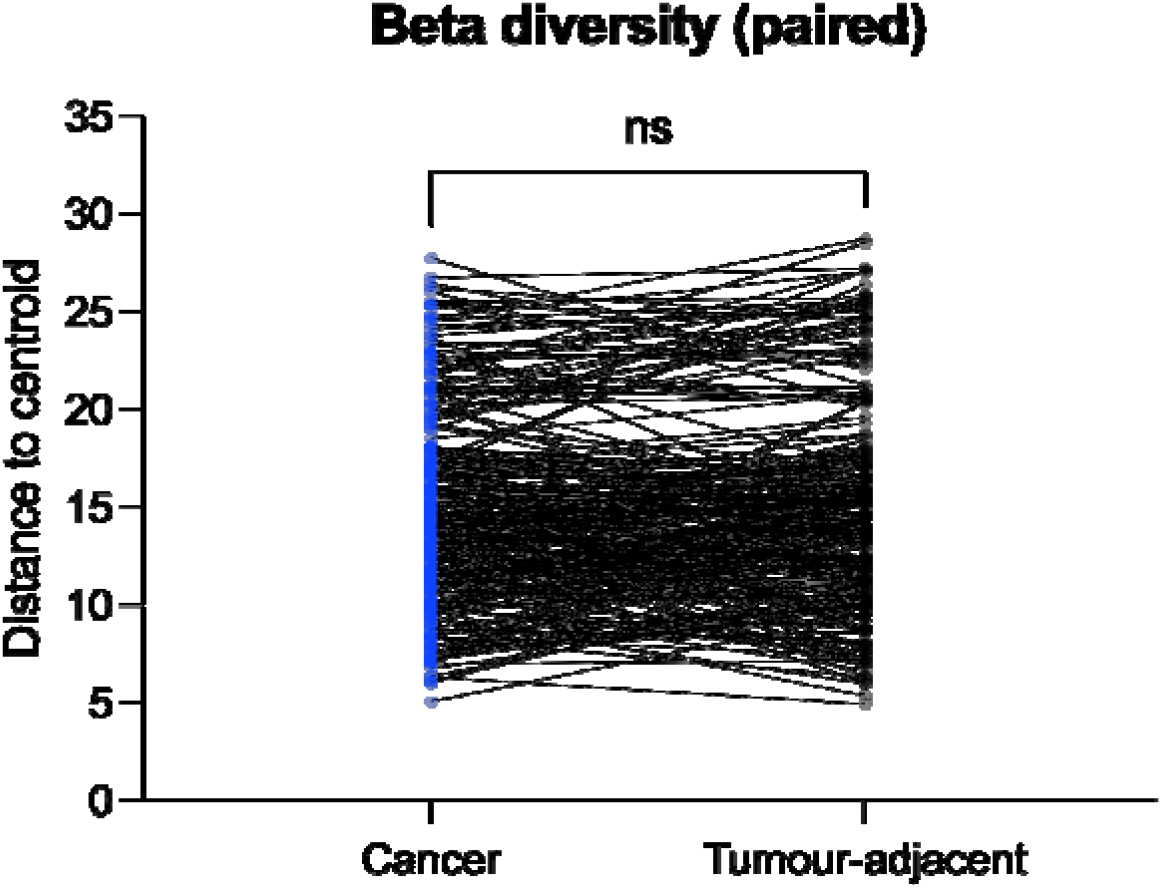
Beta-diversity between paired cancer and cancer-adjacent tissue samples. Paired Wilcoxon test was performed on Euclidean distance between each samples. No significant differences between cancer and cancer-adjacent tissue samples.

